# A mixed-methods realist evaluation of the Sutton Parenting Offer (universal parenting programme): A study protocol

**DOI:** 10.64898/2026.03.30.26348215

**Authors:** Veronica Varela-Mato, Denis M. Ngina, Elizabeth Orton, Jo Barnes

**Author notes:** Corresponding author: Dr. Veronica Varela-Mato, School of Sport, Exercise & Health Sciences, Loughborough University, Loughborough, UK. **Ethical oversight and approvals:** This study is categorised as a service evaluation and therefore does not require Health Research Authority (HRA) approval. The study has been approved by the Loughborough University Research Ethics Committee.

## Abstract

**Background:** Parenting practices shape children’s emotional, social, and cognitive developmental wellbeing. Yet, many families face complex challenges that increase the risk of poor outcomes and demand on social care. The Sutton Parenting Offer (SPO) is a universal, peer-led parenting offer that provides early, non-stigmatising support to families with children aged 0-25 years. It combines evidence-based programmes with informal workshops and peer networks delivered through Family Hubs. This present study is an evaluation protocol of the parenting offer.

**Aim:** This evaluation aims to explore how, why, and in what contexts SPO supports families in engaging, sustaining positive change, and generating wider system value.

**Methods:** A mixed-methods realist evaluation approach will be used to evaluate SPO across four work packages: engagement pathways, early changes and peer-led ecosystems, long-term change, and system value for money. Data sources will include attendance data (anonymised service records), survey data (entry and exit), and qualitative data (dyad interviews, story circles, and stakeholder-value mapping workshops). The COM-B and the Theoretical Domains Framework (TDF) will guide the analysis of behavioural data. Quantitative data will be analysed descriptively and using paired parametric and non-parametric tests, while qualitative data will be analysed thematically following a realist-informed approach to refine context-mechanism-outcome (CMO) configurations.

**Discussion:** This protocol presents the first realist informed evaluation of a universal parenting program in a local authority setting. The evaluation will generate evidence on how, when and why a universal, community-based, and peer-led model such as the Sutton Parenting Offer engages families and generates change. The findings will be useful to inform future parenting service design and implementation in local contexts in England.

## BACKGROUND

Healthy and safe parenting underpins children’s social, emotional, and developmental wellbeing. Parenting practices have been associated with children’s emotional outcomes and childhood disruptive behaviours (1,2). These behaviours are in turn associated with a range of adverse outcomes, including lower educational attainment, criminal involvement, unemployment, and social exclusion (1).

Nationally, 4.6% of children aged 5–19 meet criteria for behavioural disorders (3), one of the most common reasons for referral to children’s social care. Relationship distress further compounds these risks, with one in ten children living with both parents residing in households where at least one parent reports relationship difficulties (4).

Each day in the UK, around 1,700 children are referred to children’s social care, and one in five enter care due to family dysfunction, often associated with parenting practices (5). In Sutton, 2,070 children were referred to social services in 2024, with 664 starting Child in Need plans, which provide additional support to help children achieve or maintain a reasonable standard of health or development (6). Currently, nearly 84,000 children are looked after by local authorities in England (7). The escalating cost of children’s social care is one of the greatest pressures facing councils, with budgets rising by 11% (£14.2 billion) in 2024/25 (5). Residential placements (children’s homes run by local authorities) cost nearly £5,000 per week (8), illustrating the substantial costs associated with specialist provision.

National policy recognises these challenges through initiatives such as the Reducing Parental Conflict programme, Family Hubs and Start for Life, which aim to strengthen family relationships and improve outcomes. Since Allen’s (2011) review, *Early Intervention: Next Steps* (9), there has been growing emphasis on early intervention, alongside the establishment of the Early Intervention Foundation to promote evidence-informed approaches (10). Parenting programmes grounded in social learning and attachment theories have been shown to improve parenting and child outcomes (2,11). However, access remains unequal. Families from disadvantaged backgrounds are less likely to participate, and support is often delivered reactively to families considered at high-risk rather than offered universally (12).

By embedding evidence-based programmes within a universal, peer-led, and community-rooted structure, the Sutton Parenting Offer (SPO) seeks to provide non-stigmatising support to all families with children aged 0–25 years. The service combines structured evidence-based programmes, including Empowering Parents, Empowering Communities (EPEC), Solihull Online, and Circle of Security, with informal workshops, and peer-led groups. It also offers targeted provision for specific needs, such as teenage parenting, additional needs, and father engagement.

Delivered across schools, community venues, and family hubs, SPO adopts a “no wrong door” model to ensure accessible, inclusive, and culturally relevant support for diverse families. Through this approach, the service aims to improve parenting practices, strengthen family relationships, and reduce reliance on crisis services (**Figure 1**).

The present evaluation aims to address the primary research question, how, why, and in what contexts does the parenting service support families to engage and sustain positive change. To address this aim, the evaluation is structured in 4 work packages aligned to secondary research questions: WP1, Pathways to engagement; WP2, Early change and building peer ecosystems; WP3, Long-term change and breaking the cycle; WP4, System and value for money, as outlined in **Table 1**.

## METHODS

A mixed-methods realist evaluation will be conducted to explore how, why, and in what contexts SPO works. Complex public health services such as SPO involve multiple interacting components delivered across varied settings (**Figure 1**), making their implementation and impact highly context dependent (13). Realist evaluation is a theory-driven approach that explains how outcomes are generated by examining the interaction between context, mechanism, and outcome (CMO) configurations (14). It involves developing and refining theories of change (ToCs) that describe the pathways through which services are expected to achieve impact. Rather than focusing solely on whether an intervention works, realist evaluation seeks to understand how specific contexts enable or constrain mechanisms of change (14).

The realist approach is methodologically neutral, drawing on diverse data sources to iteratively test and refine programme theories (15). This flexibility makes it particularly well-suited to evaluating complex, multi-level, and adaptive public health interventions. Realist methods have been used to assess interventions in areas such as mental health, smoking cessation, physical activity, and parenting support, as well as in local authority services addressing food advertising policy and peer parental advocacy (16,17). Building on this knowledge, the present evaluation will develop and refine explanatory ToCs that capture how the SPO generates outcomes across different contexts.

### 1. Development of initial ToCs

Initial ToCs were co-developed through extensive co-production and participatory workshops involving end-users, frontline practitioners, and senior leadership team (SLT) members. These sessions captured diverse perspectives and narratives illustrating how the service, or specific components, may generate change in different contexts.

In parallel, a review of peer-reviewed literature identified mechanisms and outcomes reported in comparable interventions. Together, these activities informed the development of 56 preliminary CMO configurations.

Theory consolidation involved systematically reviewing, clustering, and refining these preliminary CMOs. Similar or overlapping configurations were grouped thematically, with attention to recurring mechanisms such as trust-building, peer validation, and relational safety and the contexts in which they were activated.

Through this process, the CMOs were synthesised into four overarching ToCs, aligned with short-, medium-, and long-term goals and value for money considerations. **Table 2** presents these ToCs and their alignment with the work packages.

#### Data collection

Data will be collected using different methods across the four work packages to explore different aspects of family engagement, peer-led support, long-term change, and overall service value. Table 3 outlines the data collection plan, including the methods, sample sizes, target populations, recruitment timelines, and the purpose of each activity. A combination of different methods will provide a comprehensive understanding of how families interact with the service, the changes they experience over time, and the value created for families, community, and local systems.

##### a) Service data

Anonymised sociodemographic data will be extracted from the Early Start (EStart) Database, a secure local authority system recording family-level information across early help, parenting, and wellbeing services. Variables will include age, gender, ethnicity, spoken language, Special Educational Needs (SEN) and IMD.

Where available, developmental and educational indicators, including Early Years Foundation Stage Profile (EYFSP) outcomes (Good Level of Development, Early Learning Goals) and Attainment 8 scores (a measure used in England to assess a pupil’s average achievement across eight *key* qualifications at the end of secondary school) will contextualise family needs and outcomes. Referral patterns and service requests will provide insight into pathways to engagement.

Social care tiers from the Mosaic database will describe changes in support levels, such as transitions from Child in Need (CIN) to Early Help or case closure. Data will be analysed descriptively to summarise family characteristics and engagement patterns, with comparisons across key demographic variables including ethnicity, IMD, and parent gender.

##### b) Qualitative data and survey data

In addition to routinely collected data, new knowledge will be generated through a survey and qualitative methods as presented in **Table 3**.

The COM-B model provides a concise structure comprising Capability, Opportunity, and Motivation, which interact to drive behaviour change, which in this context is improvements in parenting practices. The TDF complements COM-B by offering greater specificity through 14 domains, including knowledge, skills, social influences, beliefs about capabilities, and environmental context. Used in tandem, these frameworks enable a multi-level analysis of behaviour determinants. COM-B provides an overarching structure, while the TDF supports systematic identification of specific barriers and facilitators to implementation.

#### Study population

The participants will comprise parents or carers and children who meet the following criteria.

##### Parents or carers

Eligible parents or carers will:

- Be primary carers, including parents or legal guardians in community and prison, of children and young people aged 0 to 25 years.
- Be aged 18 years or older with no upper age limit.
- Be able to provide informed consent.
- Be residents of Sutton

Parents or guardians under the age of 18 will not be included.

##### Children

Children participating in the creative workshops will:

- Be those currently supported by the SPO.
- Be aged over 5 years and under 18 years.
- Have parental consent provided.
- Be able to give age-appropriate assent if under 16 years or informed consent if aged 16 to 18 years.

The following children will not be included:

- Children aged 18 years or under who have their own children.
- Children for whom parental consent is not provided.
- Children in social care

Participation will also not proceed where involvement is assessed as not being in the child’s best interests by LBS staff or the research team due to safeguarding or wellbeing concerns.

#### Sampling strategy

##### Qualitative data

The sample sizes for qualitative data collection in this study are informed by established literature on qualitative research and realist evaluation. Discussion groups generally produce stable findings with four to eight groups of 5–7 participants (18). Dyadic interviews, in which two participants share experiences together, typically require 12–16 cases to capture relational dynamics (19).

Recruitment will continue until analytic saturation (20), is reached. Interviews and focus groups will be transcribed and analysed in batches to monitor the emergence of new themes (21). Where fewer than 5% of themes are novel, additional data collection will be considered unlikely to generate substantial new insights.

This approach aligns with contemporary perspectives emphasising data adequacy, whereby the dataset is judged sufficiently rich and credible to address the study objectives (20,22). This approach supports the generation of trustworthy findings while minimising participant burden.

##### Quantitative data

###### Survey

A convenience sample will be used. Approximately 60 families are expected to enrol across the various service cohorts during the data collection period, from October 2025 to March 2026. Drawing on literature related to the involvement of vulnerable groups in research (23,24), an attrition rate of 40–50% is anticipated.

##### Service data

The sample will include all families who engaged with the service, from the first cohort in December 2021 to the final cohort in March 2025 (approximately 1,000 families). Children’s test results from the March 2025 cohort will be included in the analysis, regardless of the timing of completion.

#### Data analysis

##### Qualitative data analysis plan

Interviews and discussion groups will be audio-recorded and transcribed verbatim. Transcript will be checked against audio files to ensure accuracy. All identifying information will be removed, and participants assigned unique identifiers to maintain pseudonymity. Qualitative data will be managed using NVivo and analysed using a realist-informed thematic approach.

Coding will adopt a hybrid approach. Deductive codes will be informed by the COM-B model, the TDF, and the initial theories of change. Inductive codes will capture emergent concepts grounded in participants’ accounts. Codes and subcodes will be organised into themes, which will then be integrated and configured into provisional context–mechanism–outcome (CMO) explanations.

To support systematic exploration and data adequacy, transcripts will be analysed in small batches. After each batch, newly identified codes and themes will be reviewed and compared with existing analytical categories to assess whether additional explanatory insights are emerging. This iterative process will continue until no substantively new mechanisms or contextual insights are identified, indicating sufficient analytic depth. One researcher will conduct initial coding and framework development. To enhance analytical rigour, a second researcher will independently code 20% of the transcripts and compare interpretations. Discrepancies will be discussed and resolved through consensus, and the coding framework refined accordingly.

A reflexive approach will be maintained throughout. Researchers will document assumptions, positionality, and potential biases in reflective memos. Regular analytic meetings will provide opportunities to examine these reflections and challenge emerging interpretations.

An audit trail will document coding decisions, theme development, and revisions to the CMO configurations. This will include NVivo memos, analytical notes, meeting summaries and versioned coding frameworks to support transparency and methodological integrity.

Integration will occur through triangulation across data sources, including parent and facilitator interviews, discussion groups, observational fieldnotes, and open-ended survey responses. Patterns of convergence and divergence will be examined to identify recurring mechanisms and contextual influences.

Provisional CMO configurations will be iteratively refined through comparison with initial theories of change and mapped against relevant COM-B and TDF domains. Emerging explanations will be discussed during team-based analysis sessions and explored in member-checking workshops with interest holders to enhance plausibility and practical resonance.

Findings will be reported through thematic summaries, refined CMO tables, and visual logic models illustrating explanatory pathways. Illustrative excerpts will be included to demonstrate the grounding of interpretations while preservice participant’s voices.

##### Quantitative data

Descriptive statistics will be used to summarise the characteristics of the study sample and key variables. Categorical variables will be reported as frequencies and percentages. Continuous variables will be assessed for normality and summarised using means and standard deviations if normally distributed, or medians and interquartile ranges if skewed.

Changes in between entry and exit surveys will be analysed using paired t-tests or Wilcoxon Signed-Rank Tests, as appropriate. To assess potential attrition bias, demographic characteristics and baseline scores (confidence, stress, wellbeing) will be compared between completers (participants with both entry and exit data) and non-completers using Chi-square tests for categorical variables and independent samples t-tests or Mann–Whitney U tests for continuous variables, as appropriate.

Relationships between categorical variables will be examined using Chi-square tests. Comparisons of continuous variables across multiple independent groups will use one-way ANOVA for normally distributed data or Kruskal-Wallis tests for non-normally distributed data. Ordinal variables will be analysed using Kruskal-Wallis tests.

Context-specific items not repeated across surveys (e.g. questions about first contact experiences or perceptions of inclusivity) will be presented descriptively using frequencies and percentages.

#### Triangulation

Triangulation will be applied at three levels to strengthen the robustness and credibility of the evaluation findings.

### 1. Data triangulation

Data will be collected from multiple sources and at different stages of the evaluation to answer the research questions (Table 1). This will include perspectives from parents, peer facilitators, service providers, and local authority staff, alongside survey responses and service monitoring data. Collecting data across different participant groups and time points will enable comparison of experiences and contexts, supporting the refinement of CMO configurations.

### 2. Methodological triangulation

The evaluation will combine qualitative methods (interviews, discussion groups, creative workshops, observations, open-text survey responses) with quantitative approaches (survey data and service monitoring data). Secondary data sources will also be drawn upon where appropriate. Using multiple methods iteratively across the work packages will enable the evaluation to explore both breadth (patterns and trends) and depth (narratives and mechanisms), and to test how quantitative evidence aligns with or diverges from qualitative accounts.

### 3. Researcher triangulation

Multiple study researchers will be involved in analysis to minimise bias and strengthen rigour. An experienced lead researcher will provide oversight and review to mitigate observer bias. In addition, emerging findings and refinement of CMO configurations will be discussed regularly within the research team and shared formatively with PPI contributors to ensure transparency and enhance interpretation.

#### Integration and reporting

Findings from these triangulation layers will be integrated to form a cohesive exploratory narrative. This integration will allow cross-verification of results, highlight convergence and divergence between different perspectives, and reduce the likelihood of bias. Triangulated findings will provide a comprehensive understanding of:

How, why, and in what contexts does the Sutton Parenting Offer to support families to engage and sustain positive change.

1. How does the SPO generate change.
2. The barriers and facilitators influencing engagement, behaviour adoption, and the role of peer ecosystems within the support system.
3. The short- and longer-term impacts on parents’ knowledge, skills, and wellbeing.
4. How, why, and for whom (families, communities, and the local authority) the service creates value, and under what conditions this value is sustained or diminished.

#### Public involvement

The evaluation has been co-designed by the study team in collaboration with the London Borough of Sutton, local partners, stakeholders, and end-users. A multi-stakeholder working group was established early in the process, comprising the research team, a public contributor, senior leadership representatives, and frontline practitioners. Through regular meetings, online sessions, and in-person creative workshops, including story circles and LEGO® SERIOUS PLAY® with parents and online stakeholder discussions, public and professional perspectives were used to shape the evaluation approach. These activities focused on capturing experiences of how the service supports parents and on co-designing the research questions and methods, leading to the development and refinement of a shared logic model and agreement on the evaluation aims and primary research questions. The research plan outlined in this protocol reflects an extensive process of needs assessment and prioritisation, grounded in lived experience. Public involvement will continue through a local PPI group, supported by the study PPI lead and a member of the Public Advisory Group, with public contributors working alongside researchers to review data collection tools, support data collection, contribute to analysis and interpretation, and co-produce accessible dissemination outputs.

## DISCUSSION

### Study rationale and contribution

This protocol outlines a realist evaluation designed to understand how and in what contexts a universal, peer-led parenting service operates within a local authority setting. Although substantial evidence demonstrates that structured and peer-led parenting interventions can improve child behaviour, parental wellbeing, and engagement, particularly in disadvantaged communities, much of this literature emphasises outcomes rather than the mechanisms and contextual conditions through which change occurs (2).

Programmes such as Triple P and EPEC demonstrate positive impacts on parenting practices and child outcomes. However, less attention has been paid to how universal, community-based parenting services function in real-world systems, particularly in relation to engagement, sustained participation, and perceived value at both individual and system levels. This evaluation addresses that gap by generating explanatory evidence of how the SPO operates within its local context, rather than testing effectiveness.

### Use of a realist, theory-driven approach

A realist framework is well suited to capturing the complexity and contextual sensitivity of SPO. Parenting services delivered within local authority and community settings are shaped by interacting factors, including delivery models, facilitator roles, family circumstances, and wider system constraints (25), which are not adequately addressed by evaluations focused solely on outcomes.

This study adopts a theory-driven approach in which initial programme theories were co-developed with parents, practitioners, and system leaders and will be refined iteratively through empirical data. The mixed-methods design enables systematic examination of how contextual conditions activate mechanisms to generate outcomes, supporting explanatory insight into how the service works for different families.

### Universal and non-stigmatising support

SPO is delivered as a universal service, contrasting with targeted interventions that may inadvertently reinforce stigma or deter engagement. Evidence suggests that universal and group-based approaches can enhance accessibility and reduce stigma, particularly for families in disadvantaged or marginalised communities (26,27).

This evaluation will examine how universal access, community embedding, and non-stigmatising delivery shape pathways into the service and patterns of participation.

Rather than assessing effectiveness, the study seeks to understand how universal models operate within local authority systems and how value is experienced by families and providers.

### Peer-led delivery and community-rooted practice

Peer leadership is central to SPO and is hypothesised to operate through mechanisms such as trust, shared experience, and relational safety. Evidence indicates that peer-led, group-based parenting programmes can successfully engage socioeconomically disadvantaged communities (2,26,27), where shared learning and reduced power differentials may strengthen engagement.

This evaluation will explore how peer-led elements interact with structured programmes and informal provision across contexts, and how these interactions shape perceptions of relevance, accessibility, and sustained engagement. It will also examine whether peer-led delivery mitigates barriers to participation among underrepresented groups, including fathers.

### Early intervention and system relevance

The evaluation aligns with national priorities on early intervention, Family Hubs, and reducing demand on statutory children’s services. Parenting interventions are increasingly positioned as core components of early help systems, yet evidence remains limited regarding the contribution of universal, community-based services to early intervention pathways in practice.

This study will examine how SPO is perceived by families and professionals to support early help and how it integrates within the wider local system. System value will be explored through stakeholder accounts of how peer-led and community-embedded mechanisms may contribute to earlier support, reduced escalation to statutory services, and longer-term wellbeing and economic value, without undertaking formal cost-effectiveness modelling.

### Anticipated limitations

As a service evaluation, findings will be context specific and are not intended to provide generalisable estimates of effectiveness. The study relies partly on routinely collected service data, which may vary in completeness and consistency. Attrition is anticipated in survey and qualitative components due to the voluntary nature of participation and the characteristics of the population.

These limitations are inherent in real-world evaluations of community services. However, the realist approach supports analytical transferability by identifying mechanisms and contextual conditions that may be relevant to similar settings, rather than seeking statistical generalisation.

### Implications and future directions

Findings will inform service design and commissioning decisions, particularly in relation to early intervention, peer leadership, and integration within Family Hub and early help systems. Insights into engagement processes, trust-building, and perceived system value will support decisions about service adaptation, sustainability, and potential scale.

For the research community, the study contributes methodological learning on applying realist evaluation to universal, community-based services. The refined theories of change will provide a foundation for future comparative research, longitudinal follow-up, and formal economic evaluation.

## Conclusion

This protocol presents a transparent and theory-driven evaluation of the Sutton Parenting Offer, a universal, peer-led parenting service delivered within a local authority context. By articulating the evaluation framework, analytic strategy, and theoretical underpinnings in advance, the study enhances methodological rigour and transparency. It will generate locally relevant explanatory evidence to inform service development while contributing to wider debates on evaluating complex, community-based parenting support within public systems.

## Supporting information

SPO supplemental file

## Data Availability

All data produced in the present work are contained in the manuscript

## Abbreviations

CMO: Context Mechanism Outcome
COM-B: Capability, Opportunity, Motivation, and Behaviour
EPEC: Empowering Parents, Empowering Communities
LBS: London Borough of Sutton
PPI: Patient and public involvement
SPO: Sutton Parenting Offer
TDF: Theoretical Domains Framework
ToC: Theory of Change
WP: Work Package

## Acknowledgements

We acknowledge Sarah Alderman, Laura Devereux, Theresa Cameron, and Claire Borg from the London Borough of Sutton for their contributions to the design of this evaluation. We also thank Swati Madhavji, Public Contributor Group Lead, and all parent contributors whose insights were central to shaping this research protocol.

## Declarations

NA

## Consent for publications

NA

