## Supplementary material for "A mixed-methods realist evaluation of the Sutton Parenting Offer (universal parenting programme): A study protocol": SPO supplemental file

**Table 1: Research questions and aim per work package**

| ***Primary Research Question -*** *How, why, and in what contexts does the parenting service support families to engage and sustain positive change?* | | |
| --- | --- | --- |
| ***Work Package*** | ***Aim*** | ***Secondary Research Questions*** |
| *WP1. Pathways to engagement* | To explore how and *why* families, particularly those from underserved or marginalised groups, decide to engage (or not) with peer-led parenting support, focusing on how early experiences of trust, warmth, and recognition of shared experience influence motivation and willingness to participate. | In what contexts do families choose to engage (or not) with the service, and *why* do mechanisms such as trust, relational safety, peer support, and recognition of “people like me” shape these decisions? |
| *WP2: Early change and building peer ecosystems* | To examine how and *why* mechanisms such as validation, relational safety, and peer-led support enable parents to build skills, knowledge, and participation, and how these early relational and reflective changes form the basis for sustained confidence, emotional connection, and behavioural change for families over time. | In what contexts do parents experience early changes in skills, knowledge, participation, and peer support, and why do mechanisms like validation, relational safety, and the growth of peer ecosystems bring about these outcomes? |
| *WP3: Long-term change and breaking the cycle* | To examine *how and why* emotionally safe, trauma-informed, and peer-led support enables families to reflect on their past, rebuild trust, and sustain long-term changes in parenting practice, relationships, and emotional wellbeing.  This includes understanding the mechanisms that help parents maintain change beyond programme participation, and the contextual factors that support or hinder this sustainability. | In what contexts are families able to sustain positive changes beyond the programme, and *how and why* do mechanisms such as trauma-informed support, ongoing peer networks, and trust-building processes influence the maintenance or decline of these changes over time? |
| *WP4: System and value for money* | To examine *how, why, and under what conditions* the service generates value for families, communities, and the local authority. This includes understanding how peer-led and community-embedded mechanisms strengthen trust, prevent crisis escalation, and create longer-term wellbeing and economic benefits and why this value is sustained or lost over time. | In what contexts does the service generate value for families, communities, and the local authority, and *how and why* do peer-led and grassroots mechanisms contribute to wider system change and sustained impact? |

**Table 2: Theories of Change and work packages**

| **Work Package** | **Theory of Change** |
| --- | --- |
| WP1. Pathways to engagement | When families are welcomed in a warm, flexible, and non-judgemental way, their initial fears, stigma, or mistrust begin to shift, leading to voluntary engagement and early buy-in. This happens because relational safety and recognition of shared experience reduce defensiveness and help families feel accepted and hopeful that change is possible. |
| WP2: Early change and building peer ecosystems | When parents connect with relatable leaders in a non-hierarchical space, peer-led encouragement and shared identity foster trust, embedded parenting strategies, and leadership pathways. These mechanisms work because parents feel understood and validated by people with similar lived experiences, reducing stigma, and enabling more open reflection and learning. |
| WP3: Long-term change & breaking the cycle | When families are met with dignity, validation, and trauma-informed care, they begin to rebuild trust, reflect on their histories, and shift entrenched behaviours, laying the groundwork for healthier family dynamics and generational change. This occurs because emotionally safe and non-judgemental relationships allow families to process past experiences, restore self-worth, and experiment with new ways of relating to their children and services. |
| WP4: System and value for money | When peer-led and grassroots approaches are embedded within local authority systems, and families experience meaningful, community-rooted support, trust in services is strengthened. This enables earlier and more effective intervention, reduces reliance on costly statutory provision, and generates longer-term social and economic value for families, communities, and the local authority. This happens because families engage more readily with trusted, community-based networks, and services can respond earlier and more appropriately to need. |

**Table 3: Data collection plan**

| **Work package** | **Data source/method** | **Sample size** | **Target population** | **Recruitment** | **Purpose** |
| --- | --- | --- | --- | --- | --- |
| WP1: Pathways to Engagement & Early Impact | Entry and exit survey | ~ 60 families baseline | new parents enrolling in structured support from the Sutton Parenting Offer | Oct 2025 to March 2026 | To understand pathways into the service, reasons for engaging/not engaging, cultural safety, barriers to access, and early shifts in knowledge and skills |
|  | Service data |  | Families that have engaged with the service since December 2021 till March 2025 |  | To understand referral pathways (self-referred or internal/external services), request reason and who requested the service |
| WP2: Early change and building peer ecosystems | Dyad interviews coupled with ecosystem mapping* | 12-16 dyads | Parents mid-programme and their facilitators | Rolling recruitment Nov 2025 to Apr 2026 | To explore how peer-led support helps families feel more confident, connected, and supported in their parenting. To assess how feeling understood, safe, and part of a peer network supports early changes in families |
|  | Service data |  | Families that have engaged with the service since December 2021 till March 2025 |  | To examine transitions in support levels. Baseline status (e.g. Child In Need (CIN)) will be compared descriptively with subsequent outcomes, including step-down to early help or case closure. |
| WP3: Long-Term Change & breaking the cycle | Story Circles with service journey sketches | Four to six groups with five to seven participants each (20–42 participants) | Parents 3–6 months post-programme; peer facilitators; professionals | Rolling recruitment Nov 2025 to Apr 2026 | To explore long-term change, including sustained identity shifts, emotional safety, accountability, and the transfer of trust into wider services. To capture parents’ journeys through the service, focusing on entry points, turning points, barriers, enablers, and wider impacts such as family relationships and access to services. |
|  | Child creative sessions | Two groups of four to six children | Children and young people currently supported by the service | Rolling recruitment Nov 2025 to Apr 2026 | To understand what has changed at home or for the child since they or their family started taking part in the Sutton Parenting Offer. |
|  | Service data |  | Families that have engaged with the service since December 2021 till March 2025 |  | To describe changes in Good Level of Development and Early learning Goals. Average scores for children in families engaged with the service will be analysed descriptively and benchmarked against borough-wide and (CIN) averages. This benchmarking will provide contextual insight into relative outcomes but is not intended for causal inference. |
| WP4: Value for money and sustainability | Stakeholder Value Mapping Workshops | Two to three sessions of 8–12 participants | Local authority stakeholders, peer leaders, frontline professionals, families | Month 2–3 (scoping), Month 5 (mid), Month 8 (final). | To assess the value of the service, the research will explore alternative pathways that families might follow if the service did not exist, the associated costs, and the gaps currently filled. The analysis will also consider wider contributions, such as the impact of peer-led, community-based activity on system change. Volunteer contributions will be valued using a time-banking approach. Findings from this analysis will inform the development of a framework for the local authority to assess the social and preventative outcomes (SPO) of the service in the future. |
|  | Mini Interviews (15 min) | Four to six participants | Participants attending the value mapping workshops | Within 14 days of attending the workshop | Follow-up calls after the stakeholder workshops will be conducted to clarify specific details about how value is created, captured, and sustained within the service |
|  | Cost-data from the literature | Latest cost data on child involvement with social care | To be validated during the workshop - expert consensus | - | To understand the cost burden to Local Authority if children supported were referred to social care. |

*A participatory method used to visually map the relationships, networks, and services that surround and influence families.

**Figure 1: Logic model for evaluation**

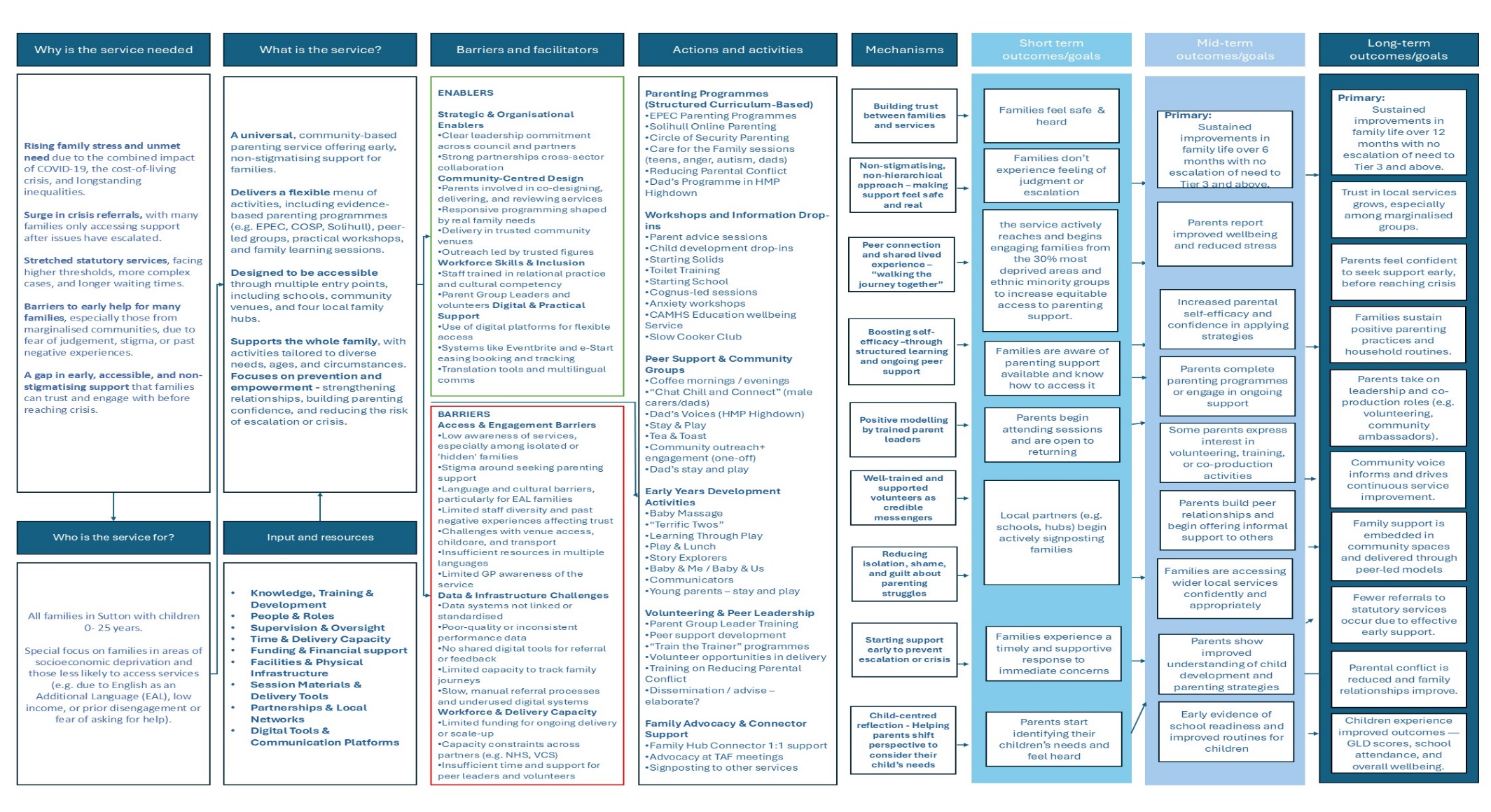
